# Evaluation and Optimization of Automated External Defibrillator Deployment: Comparing Straight-Line and Walking-Route Distances for Out-of-Hospital Cardiac Arrests

**DOI:** 10.1101/2025.05.13.25327567

**Authors:** Zhaohui Qin, Jia Li, Shuyao Zheng, Da Xu, Wei Zhang, Lu Lu, Xianliang Yan, Tie Xu, Ningjun Zhao, Yan Xu

## Abstract

**Background:** This study aimed to explore appropriate automated external defibrillator (AED) deployment strategies for Gamma cities, addressing the gap in research. By comparing the coverage of AEDs under different calculation models, we evaluated the rationality of the distribution of the AEDs in a Gamma city within eastern China and provided suggestions for improvement.

**Methods:** This observational study used two datasets (an AED inventory and an OHCA registry) to calculate the average distance between out-of-hospital cardiac arrest (OHCA) cases and the nearest AED, and assess AED coverage rates based on straight-line and walking-route models. A total of 1,350 OHCA cases and 1,238 AEDs were included.

**Results:** The average emergency radius per AED was 497.96 m. The average walking-route and straight-line distance between OHCAs and the nearest AED were 407.09 and 378.94 m, respectively. The AED coverage rate within 100 m was 8.30% and 7.33% based on the straight-line model and walking-route model, respectively. The coverage rate within the golden six minutes for resuscitation using the walking-route model was 39.26%.

**Conclusions:** The AED distribution throughout the studied city was generally well-organized, with a large number of devices within close proximity and good coverage. However, a significant standard deviation was noted, indicating a clear clustering rather than a uniform distribution. The coverage rate within the golden six minutes was moderate. However, in an urban environment, the straight-line model may underestimate the average distance and overestimate the coverage. Hence, the walking-route-based model may be more helpful considering the critical nature of OHCA.

## Introduction

Cardiac arrest (CA) is a critical health concern that severely affects human life. Among the various types of CAs, out-of-hospital cardiac arrests (OHCAs) have consistently exhibited high mortality rates over the years. According to emergency medical services (EMS) statistics from various regions throughout China, OHCAs have an incidence rate of 97.1 per 100,000, with a return of spontaneous circulation (ROSC) rate of 6% and a survival-to-discharge or survival-to-30-day rate of only 1.2% ^1,2^. This is primarily due to the development of arrhythmias (e.g., tachycardia in shock) among cardiac arrest patients as treatment time increases, which significantly decreases their survival rates ^3^. The use of automated external defibrillators (AEDs) within the golden six minutes of emergency care can significantly improve the survival outcomes of OHCA patients. In fact, international guidelines recommend that AEDs be placed in areas within 100 m of OHCA incidents, allowing bystanders easy access ^3,4,5^. Although studies indicate that the proportion of the Chinese population capable of performing cardiopulmonary resuscitation (CPR) has increased to 17.0%, usage rates of AEDs by bystanders remain below 0.1% ^1^. As such, emergency care research has focused on ensuring that OHCA patients can access AEDs for defibrillation as quickly as possible. Recently, many scholars have developed mathematical and geographic models to improve AED accessibility ^6,7^.

In Western countries and regions, AEDs are typically placed in public spaces, such as restaurants, shopping malls, and schools ^8,9,10^. However, only recently have these countries started to consider AED placement in residential communities and other high incidence OHCA zones outside public spaces ^11^. Unfortunately, AED deployment strategies utilized in Western countries may not be directly applicable to China due to difference in regional circumstances. At present, research on AED distribution in East Asia has been scarce, with studies primarily concentrated in economically developed regions, such as Hong Kong and Japan ^12,13^.

The current study focuses on AED deployment in the central urban area of a city in Eastern China. Using historical OHCA case data from this region, we aimed to examine the statistical distribution of the distances between OHCA incident locations and the nearest AEDs based on both straight-line and walking routes. This study also aimed to improve AED accessibility for OHCA patients by investigating the impact of urban environments on AED coverage. Ultimately, our results provide a scientific basis for governmental bodies involved in the development of AED placement strategies.

## Methods

This study is a secondary data analysis of OHCA cases reported by emergency medical centers in a city in Eastern China from 2021 to 2023. Descriptive analysis was conducted based on historical OHCA case data. As the city does not have a dedicated OHCA database, cases were selected through secondary screening of all cases received by the emergency medical centers. The dataset includes information on the location and time of OHCA events and was recorded through a mini-program jointly developed by the city’s emergency medical services and Qilu Medical College of Shandong University. Data are limited to the central urban areas of the city. The project is titled BASIC, “National Science & Technology Fundamental Resources Investigation Project” (2018FY100600 and 2018FY100601).

In this study, all AEDs analyzed were deployed by the city’s Health and Family Planning Commission in collaboration with Mindray, a medical technology company, and were freely available for use by all residents. Information regarding the AEDs, including their usage and locations, was sourced from Mindray’s official website.

To analyze the geographic information for both OHCA cases and AED locations, we utilized the Gaode Map coordinate system to determine the latitude and longitude of each location. Geographical data were then imported into ArcGIS software. Using the spatial analysis function, we calculated the distance between each OHCA location and the closest AED in meters. Additionally, we computed the average emergency rescue radius of the AEDs and their coverage range. City map data were obtained from the Bigemap GIS website.

OHCA geographic information system (GIS) data were also imported into ArcGIS, after which the road network system was utilized along with the “Spatial Analyst” tool to analyze the walking-route distance between each OHCA location and its nearest AED. The Haversine formula was then used to calculate straight-line distances.

We assumed that the average pedestrian walking speed was 1.8 m/s. The Gaode Map walking path planning API was used to estimate the time required for pedestrians to reach an AED. A threshold of 3 min (half the golden six minutes for emergency care) was used as the baseline to calculate the proportion of OHCA cases that could be reached within this critical time frame.

The study received approval from the city’s Health Emergency Office, and informed consent was obtained for the data use.

## Results

From 2021 to 2023, a total of 1,350 cases of out-of-hospital cardiac arrest (OHCA) occurred in the central urban area of the city. These cases were categorized based on their location, including communities, schools, medical institutions, commercial streets, government offices, highways, enterprises, train stations, and tourist attractions. The majority of OHCA events occurred within communities, accounting for 1,058 cases (78.4%). The next most common locations were commercial streets, with 93 cases (6.9%), and medical institutions, with 49 cases (3.6%). The remaining locations, such as government offices, highways, and tourist attractions, each accounted for approximately 20-30 cases (Table 1). A map showing the distribution of OHCA cases in the city’s central urban area from 2021 to 2023 is presented in Figure 1.

**Table 1.** Location of out-of-hospital cardiac arrest cases.

| Location type | Number (%) |
| --- | --- |
| Community | 1058(78.4) |
| School | 26(1.9) |
| Medical institution | 49(3.6) |
| Commercial street | 93(6.9) |
| Administration | 27(2) |
| Highway | 26(1.9) |
| Enterprise | 22(1.6) |
| Station | 29(2.2) |
| Tourist spot | 20(1.5) |

**Figure 1.**
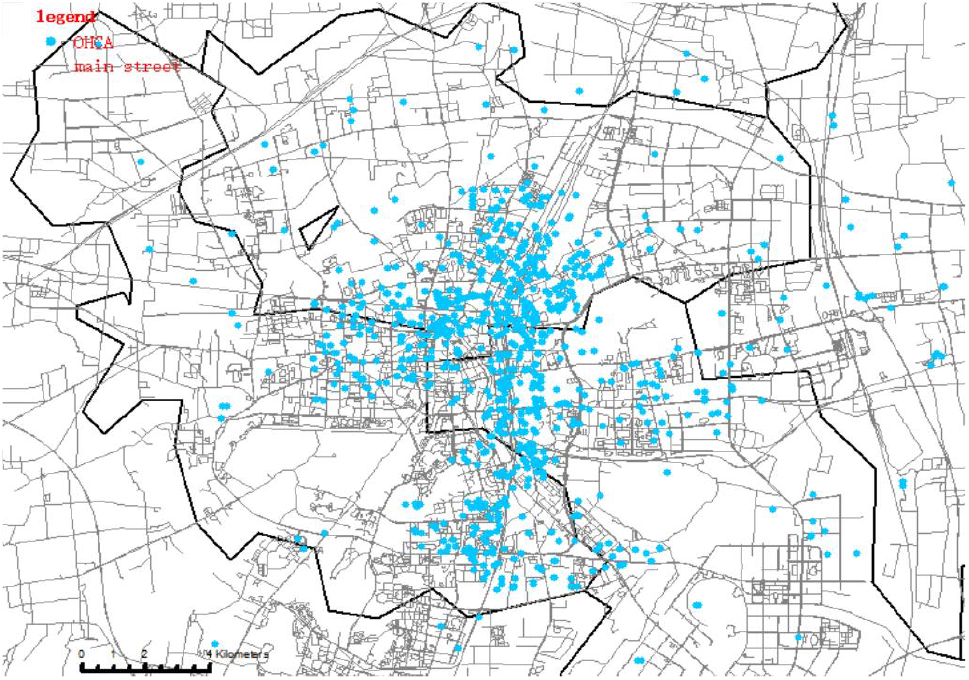
Geographic distribution of out-of-hospital cardiac arrest cases in the main urban area of the city (2021–2023)

Among the 2,968 AEDs registered on the Mindray official website during the study period, 1,238 were located in the central urban area of the city. The per capita coverage was 3.87 AEDs per 10,000 people, with a density of 1.87 AEDs per square kilometer (area data for the central urban area sourced from official government publications). AEDs were installed in various locations, including schools, communities, shopping malls, train stations, enterprises, government office buildings, hospitals, and tourist attractions. Communities (352 units, 28.4%) and hospitals (349 units, 28.2%) had the highest number of AEDs, whereas enterprises (16 units, 1.3%) and tourist attractions (15 units, 1.2%) had the least number of AEDs (Table 2). The specific distribution pattern of AEDs is illustrated in Figure 2.

**Table 2.**
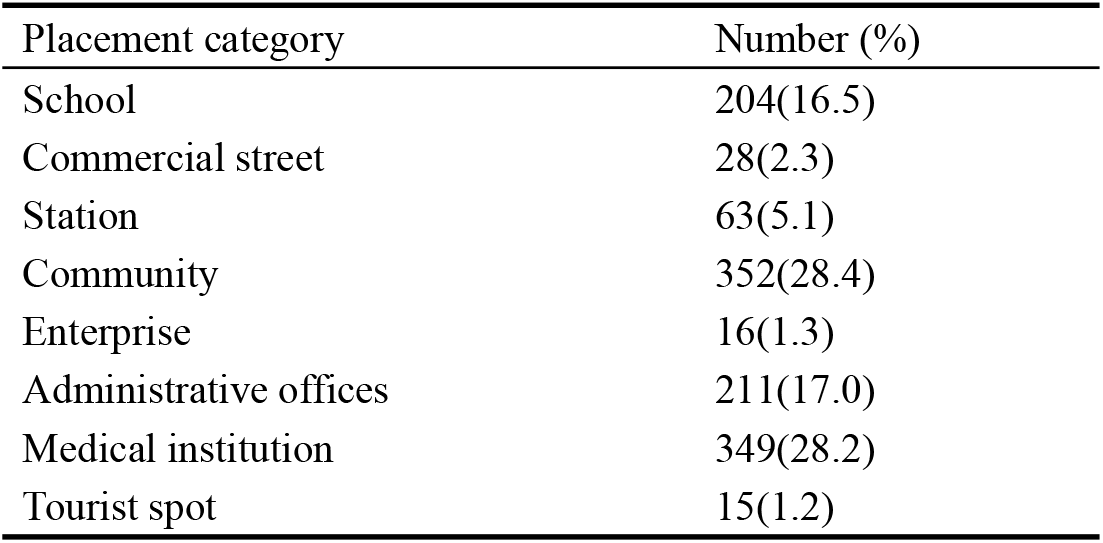
Classification of automated external defibrillator placement.

**Figure 2.**
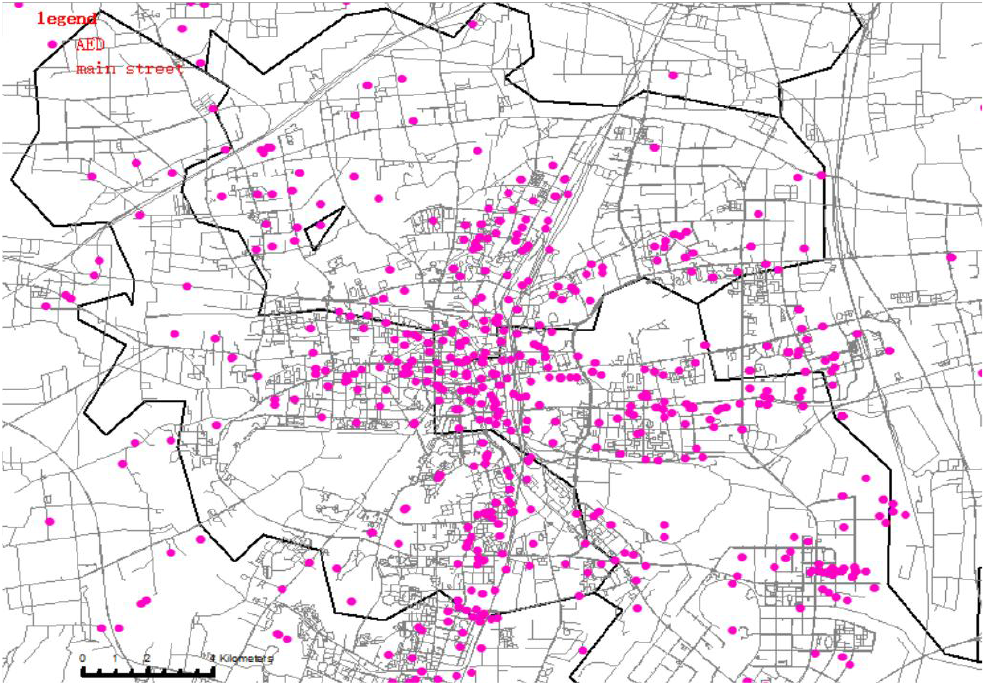
Geographic distribution map of automated external defibrillators in the main urban area of the city.

Figure 3 shows the shortest straight-line distances between AEDs. The maximum straight-line distance between two adjacent AEDs was 6,183.95 m, whereas the minimum distance was 0 m. The average distance between neighboring AEDs was 497.96 m (SD 838.17 m), with a median of 162.27 m (Table 3). As illustrated in Figure 2, AEDs in the central urban area of the city were spaced relatively closely, indicating a high density of AEDs within this region.

**Table 3.**
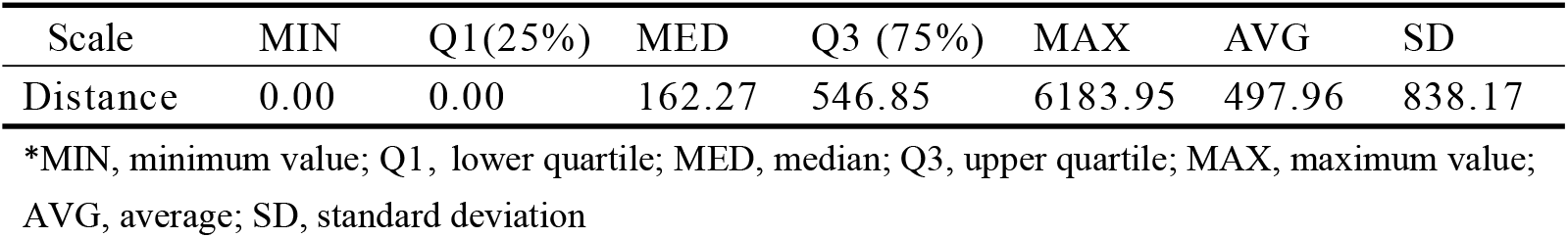
Statistical scale of the nearest straight-line distances between adjacent automated external defibrillators.

**Figure 3.**
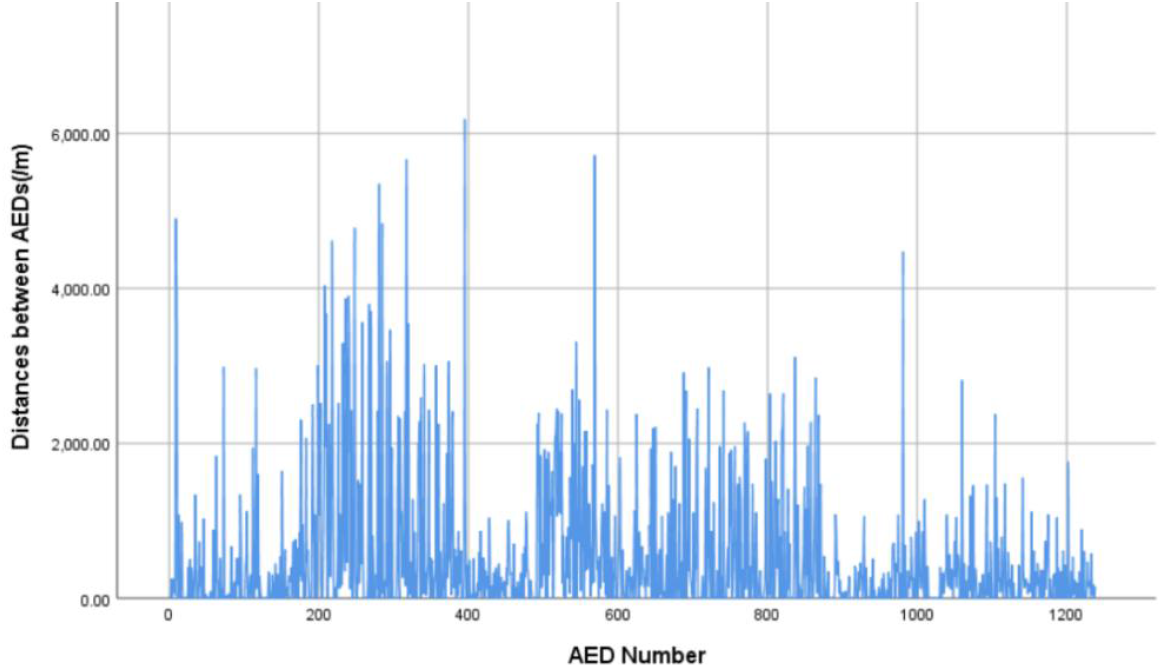
Histogram of the nearest straight-line distances between adjacent automated external defibrillators.

Figures 4 and 5 present histograms illustrating the distribution of straight-line distance and walking-route distance between the OHCAs and the nearest AEDs. The maximum straight-line distance between an OHCA case and its nearest AED was 3,993.28 m, whereas the minimum distance was 0 m. The average straight-line distance was 378.94 m (SD 351.74 m), with a median of 307.12 m. We observed a difference between the straight-line distance and walking-route distance. When converted to walking-route distance, the minimum distance remained unchanged, but the maximum distance increased to 5,343.29 m. The average walking-route distance was 407.09 m (SD 442.30 m), with a median of 327.15 m (Table 4).

**Table 4.** Distance statistics for out-of-hospital cardiac arrest cases and adjacent automated external defibrillators (/m)

| Distance | MIN | Q1(25%) | MED | Q3 (75%) | MAX | AVG | SD |
| --- | --- | --- | --- | --- | --- | --- | --- |
| Straight-Line | 0.00 | 190.75 | 307.12 | 434.31 | 3993.28 | 378.94 | 351.74 |
| Walking-Route | 0.00 | 195.06 | 327.15 | 450.00 | 5343.29 | 407.09 | 442.30 |

\*MIN, minimum value; Q1, lower quartile; MED, median; Q3, upper quartile; MAX, maximum value; AVG, average; SD, standard deviation

**Figure 4.**
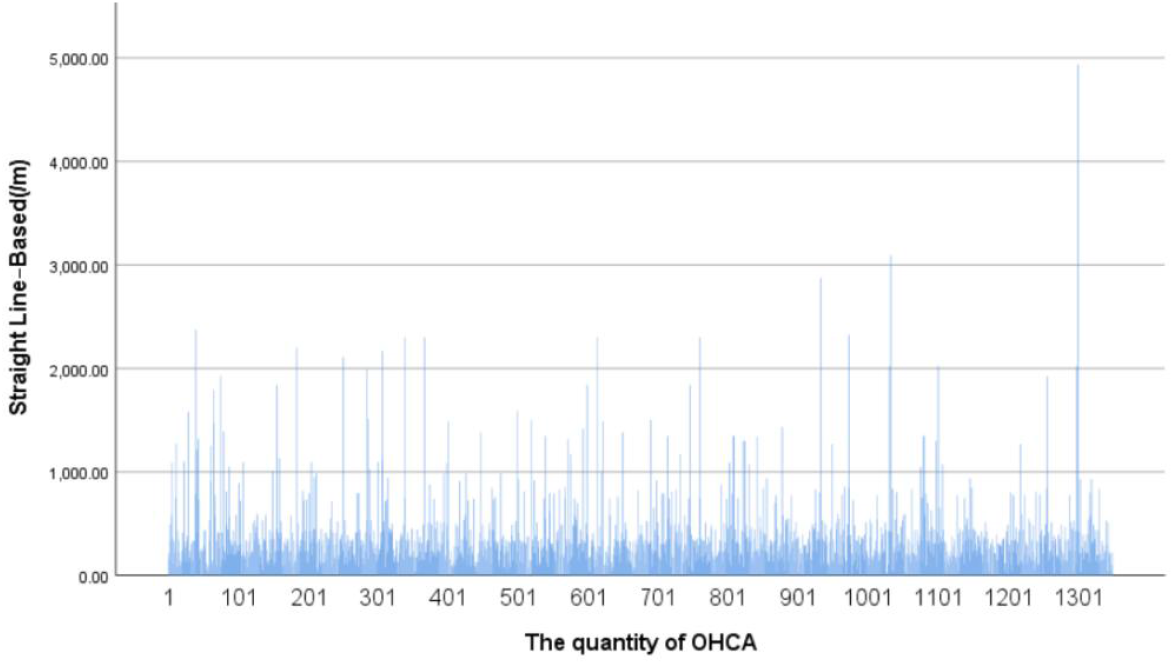
Histogram of the straight-line distance between the out-of-hospital cardiac arrest cases and the nearest automated external defibrillators.

**Figure 5.**
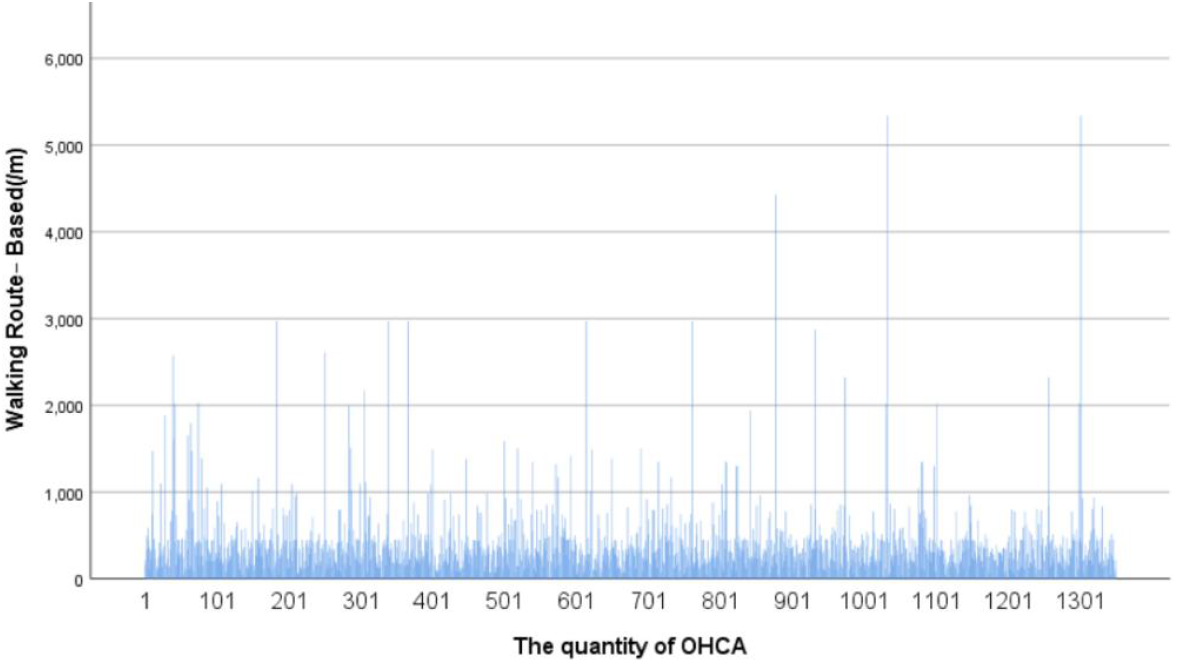
Histogram of the walking-route distance between the out-of-hospital cardiac arrest cases and the nearest automated external defibrillators.

For AED coverage, our analysis was based on the commonly used 100 m threshold for both straight-line and walking-route distances. The results showed that 112 OHCA cases (8.30%) were within a 100 m straight-line distance from the nearest AED. When converted to walking-route distance, 99 cases (7.33%) were within 100 m. Additionally, based on the threshold of half the golden hour (i.e., within 3 min), we found that 530 OHCA cases (39.26%) were reachable within the adapted walking-route distance (Table 5).

**Table 5.**
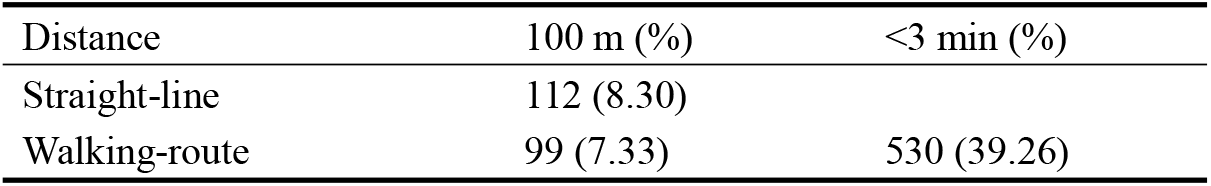
Spatial relationships between the existing automated external defibrillator and the out-of-hospital cardiac arrest.

| Distance | 100 m (%) | <3 min (%) |
| --- | --- | --- |
| Straight-line | 112 (8.30) |  |
| Walking-route | 99 (7.33) | 530 (39.26) |

## Discussion

Currently, most countries follow the guidelines for AED placement established by the European Resuscitation Council, which recommend situating AEDs in areas with high foot traffic, such as train stations, bus terminals, airports, sports facilities, and shopping centers ^14,15,16^. Our findings suggest that the AED deployment strategy in the studied city largely adheres to international conventions. The number of AEDs per square kilometer was comparable to that of Hong Kong, with the distribution of AEDs appearing to be relatively rational. However, the large SD(Standard Deviation) in placement distances indicates a concentration of AEDs in specific areas, primarily in city center administrative and commercial zones with dense populations. Conversely, industrial and agricultural areas, located farther from the city’s population centers, have fewer AEDs. The research has highlighted that cardiovascular disease mortality follows a social gradient ^17^. Individuals in underserved areas tend to experience prolonged durations of poor health, increased susceptibility to multiple diseases, and increased OHCA rates ^18^, which are further exacerbated by inequities in rural healthcare resources ^19^.

The average and median placement distances found in the current study suggest relatively high AED coverage rates within this city. In fact, the per capita AED coverage rate observed herein was higher than that in Hong Kong (1.94 per 10,000) and Busan (0.61 per 10,000) ^20,21^ but was lower than that in developed cities, such as Copenhagen (9.2 per 10,000) and Toronto (6.68 per 10,000)^22,23^.

OHCA cases collected by the city’s emergency centers were predominantly located in communities and commercial streets. GIS-based spatial relationship models between AEDs and OHCA case locations indicate that the distribution of OHCA incidents largely overlaps with the AED locations, particularly in community and healthcare settings. AED placement in these areas facilitates access for individuals in nearby communities. Although AEDs were more densely distributed in the city center than in outer areas due to the higher population density and number of OHCA incidents, our data suggest that approximately 60% of OHCA cases still face challenges in accessing AEDs within the optimal emergency response time.

Notably, our data found a relatively low number of OHCA incidents occurring in public spaces, suggesting that AED placement strategies should focus more on residential communities, particularly in and around apartment buildings. Previously, the cost-effectiveness of placing AEDs in private homes had been considered low due to high costs and inefficiency ^24^. However, the increase in AED training and emergency response education in communities may significantly improve the cost-effectiveness of placing AEDs within these areas. Recent studies also support targeting communities as an effective means of expanding health coverage for families and workers, with strategies such as placing AEDs on each floor of residential buildings showing potential for enhancing emergency response outcomes ^25,26,27,28,29^.

Despite the relatively short distances to AEDs in most residential areas, the actual use of AEDs in emergencies was far lower than the model predictions. According to data from the Mindray website, only 65 AEDs had recorded usage, despite the occurrence of 1,350 OHCA cases. The number of patients who received immediate bystander CPR was also minimal, with almost no patient receiving AED-assisted resuscitation. This discrepancy may be due to a lack of public awareness and education on the use of AEDs ^30^. The duration between OHCA onset and AED usage by bystanders often exceeds the critical 3-min threshold. Bystander response significantly influences OHCA survival rates. The duration from OHCA onset to EMS arrival involves several stages, including bystander recognition, emergency response activation, realizing the need for CPR and defibrillation, searching for an AED, and initiating rescue efforts. However, if bystanders are not aware of the need for an AED, the patient’s “chain of survival” might be interrupted ^31^.

To improve outcomes, focus should be placed not only on AED placement in high-risk areas but also on the entire survival chain, including early recognition, early bystander CPR, early defibrillation, and post-resuscitation care ^32^. A more comprehensive approach, including greater emphasis on emergency training and awareness, is necessary to support this chain and increase the effectiveness of AED deployment.

### Improvement

Given the high incidence and complex distribution of OHCA cases, establishing a unified and standardized OHCA database to facilitate more accurate tracking of epidemiological characteristics is imperative. This database should be developed based on the Utstein model, including core and optional elements such as the occurrence of OHCA, emergency dispatch data, patient demographics, treatment processes, and outcome prognoses ^33,34,35,36,37^. A standardized OHCA database would significantly enhance the scientific rigor and applicability of future studies, particularly cross-center research and comparative analyses, providing more robust and convincing evidence for clinical practice and policy-making.

Future AED deployment strategies should consider walking-route distances, especially in areas with numerous intersections where the walking-route distance may differ substantially from the straight-line distance ^13^. The disparity between the median and average placement distances in the current study suggests that multiple AEDs might be concentrated within smaller geographic areas. This finding leads one to question whether the “AEDs per 100 m^2^” metric is as effective as the widely adopted 100-m distance guideline. Despite the ubiquitous use of this 100 m^2^ threshold, its effectiveness has yet to be validated through randomized trials ^38^. Therefore, further research is needed to determine the optimal parameters for AED placement based on actual emergency response needs. We also suggested a novel model based on the “golden hour” for emergency care using the walking-route model for AED deployment, which may generate higher emergency medical efficacy in compact cities in China.

Our findings also suggest the need for expanding AED training efforts beyond specific groups, such as students, to include the general community. Emphasis should also be placed on ensuring equitable access to resources and improving national healthcare systems to address disparities in healthcare access ^39^, which should start in urban areas and gradually extend to rural, industrial, and agricultural zones. Ensuring sufficient public training on CPR and AED usage could significantly reduce delays in the chain of survival.

The widespread use of smartphones could help addressed another challenge in enhancing AED access, that is, ensuring that bystanders can easily locate the nearest AED during an OHCA incident. Accordingly, developing effective mobile applications or integrating AED location systems into existing apps could guide users to the nearest device ^40^. Moreover, some regions are experimenting with “mobile AED” initiatives, placing AEDs on taxis and buses, and training drivers to become emergency volunteers. These drivers would be notified when an OHCA occurs nearby and would deliver the AED to the scene as quickly as possible and initiate CPR and defibrillation. Pilot programs are already underway in Chinese cities like Hangzhou and Chengdu in ^41,42^. Such initiatives have the potential to expand AED coverage and improve utilization rates, ultimately enhancing public health outcomes. As technology continues to advance, new innovations offer new opportunities to improve AED access and usage ^43,44^.

### Limitations

First, the AED data used in this study were obtained from the city government’s registration records through the Mindray company’s official website. All AEDs analyzed were produced by this company and were of the same brand and model. Although the brand AED occupies most of the layout in Xuzhou city.However, AEDs purchased by other government departments or private enterprises from different brands were not included in this dataset, which may result in incomplete or skewed data.

Second, the use of AEDs may be limited by time as not all AEDs are available for use at all times. For example, AEDs may not be available for OHCA incidents occurring during nighttime hours, even with AEDs nearby, because AEDs are sometimes managed by personnel who may not be available after working hours ^45^. This time-based accessibility issue can significantly influence the actual coverage of AEDs.

Furthermore, this study assumed that the distances between AED locations and OHCA cases were measured on the same horizontal plane and only calculated the two-dimensional Euclidean distance. However, given that the city has many high-rise buildings, the actual distances between OHCA cases and AEDs could have been underestimated for AED located in or OHCA occurring on higher floors ^46^. Previous research has suggested that OHCA cases occurring in buildings may experience delays in EMS response, which further lowers the chances of survival ^13, 45^. To address this, future studies should explore three-dimensional modeling approaches, such as the Shortest Path Voronoi diagram method ^47^, which incorporates various factors, such as bridges, tunnels, and stairs that may affect walking speed, to create more precise digital models.

Lastly, the locations of OHCA incidents in this study were primarily recorded by EMS personnel accompanying ambulances. Hence, discrepancies may have been present between the recorded location and the actual site of the event, which could have also affected the true coverage of AEDs for OHCA cases.

### Conclusions

Based on the results of our analyses, the following conclusions can be formulated. First, AED placement should focus more on community areas. Second, the effectiveness of AEDs deployment based on straight-line distances from OHCA cases on a map was lower than that based on walking distances. We also can arrange AED in China within the”golden hour”for emergency care using the walking-route model. Third, despite the widespread distribution of AED, usage rates remain low. Therefore, promoting emergency education and training in communities is essential for enabling bystanders to quickly recognize the need and use of AEDs.

## Data Availability

Data Availability: All relevant data are within the paper and its Supporting Information files.

## Acknowledgments

The authors wish to acknowledge the provision of the automated external defibrillator network (https://aed-alert.mindray.com/Main/Home) by the Public Health Emergency Office of the Xuzhou Municipal Health Commission and Shenzhen Mindray Bio-Medical Electronics Co., Ltd., which facilitated access to registered AED data. The authors also express gratitude for the emergency patient database provided by the Xuzhou Municipal Emergency Medical Center, which enabled the successful acquisition of out-of-hospital cardiac arrest data.

## Sources of Funding

This study was supported by the University-Industry Collaborative Education Program by the Ministry of Education of the People’s Republic of China (231103371101213) and the Key Project of Excellent Project of Social Science Application Research in Jiangsu Province (23SYA-017).

## Disclosures

None.

## CLINICAL PERSPECTIVE

**What Is New?**

- This study conducted a systematic exploration of automated external defibrillator (AED) deployment in Gamma cities within the Chinese mainland.
- The straight-line model tends to overrate the coverage rates between an out-of-hospital cardiac arrest and the closest AED comparing with the walking-route model.
- We proposed a novel model based on the golden six minutes for resuscitation using the walking-route model for AED deployment in China’s urban context.

**What Are the Clinical Implications?**

- Strategic AED deployment in high-risk OHCA areas can improve resuscitation efficiency and optimize public health resource allocation.
- The optimized deployment of AEDs, coupled with enhanced community first-aid literacy, can improve survival rates of OHCA.

## Nonstandard Abbreviations and Acronyms

AED: automated external defibrillator
CA: cardiac arrest
CPR: cardiopulmonary resuscitation
EMS: emergency medical services
GIS: geographic information system
OHCA: out-of-hospital cardiac arrest
ROSC: return of spontaneous circulation

## Notes

### Competing Interest Statement

The authors have declared no competing interest.

### Clinical Trial

This study was conducted exclusively through computational geospatial information technology, and no human or animal subjects were involved in any phase of the research.

### Author Declarations

IRB, Xuzhou Medical University, China

